# Detecting epidemic outbreaks with routine blood tests: A comparison of surveillance methods using COVID-19 simulations and real-world seasonal disease data

**DOI:** 10.64898/2026.10.27.26364498

**Authors:** Konstantin A. Klochkov, Irina I. Maslova, Elena N. Ilina, Alexander I. Manolov

## Abstract

Timely detection of an epidemic outbreak is essential for implementing interventions and limiting the spread of the pathogen. The digitalization of healthcare has led to the availability of up-to-date, large-scale data on routinely tested blood parameters. This data can be used to detect the onset of an epidemic, which is particularly important when a new, previously unknown pathogen emerges for which no testing systems are available. In this study, we evaluated the effectiveness of various mathematical approaches for detecting an epidemic based on routinely collected blood test results. The evaluation was performed using both simulated and real-world data. First, we developed the BloodCovasim agent-based model, which defines the dependency of C-reactive protein (CRP) on time for each individual based on the course of their disease and simulates blood analysis. Using synthetic data from thousands of BloodCovasim simulations, we determined the characteristics of different methods of epidemic outbreak detection under controlled conditions. Second, we used the same methods but applied them to real-world data on CRP from Russia during a period of increased seasonal infectious diseases (rhinoviruses, adenoviruses, seasonal coronaviruses) associated with the back-to-school effect. Third, we compared the CRP-based detection method with methods based on a set of inflammatory biomarkers obtained from complete blood count (CBC) results. As a result, we found that the applicability of the methods depends on the volume of incoming data. With large amounts of data, they demonstrate similar results in terms of the probability of detecting an outbreak. As the number of tests decreases, detection methods based on basic statistical tests lose accuracy faster than those based on moving averages, while the AIC-based detector performs best. The use of composite biomarkers increases the probability of detecting an outbreak compared to using CRP alone. These results can help epidemiologists select mathematical approaches based on the volume of incoming data.

**Author summary:** To respond to an epidemic in a timely manner, it is important to detect it early. The traditional approach relies on data on cases and mortality for a specific disease, but these metrics lag behind the actual epidemic situation and require specific tests which may be absent in a case of emerging disease. Routine blood tests (e.g., C-reactive protein, CRP) can be automatically collected to accelerate the detection of an outbreak, however, the methodology for this has been insufficiently studied. In this work, we compared methods based on basic statistical tests and on trend analysis. We developed the BloodCovasim agent-based model, which simulates CRP levels during the COVID-19 pandemic. We also utilized real-world data from Russia during the period of seasonal increase in infectious diseases. We found that the applicability of different detection methods depends on the volume of incoming data. With large data volumes, the detection methods demonstrate similar performance, however, a lack of data has a much smaller impact on trend-based methods. We also found that the use of other inflammatory biomarkers outperforms CRP. The results of our work can be used by epidemiologists when selecting approaches to outbreak detection depending on the volume of incoming data.

## Introduction

Early detection of disease outbreaks is crucial for timely response to a new pathogen, effectively controlling its spread, and reducing mortality from it [1, 2]. Timely detection allows for the procurement and distribution of vaccines before an epidemic gets out of control. Traditionally, passive reporting data are used to identify epidemics, in which there is no active case finding for a specific disease [3, 4]. Such data typically include case numbers, incidence rates, cause-specific deaths, mortality rates, and counts of specific laboratory results [3, 5, 6]. This approach tends to have low sensitivity and is prone to delays [2–4, 7]. Countries are reluctant to report data due to fears of economic consequences resulting from potential restrictions on tourism and trade [3]. Underestimation of the number of cases arises from under-ascertainment (some people do not seek medical care, especially in cases of mild illness) and underreporting (some cases are not included in the statistics due to diagnostic errors or a lack of laboratory confirmation) [4, 7]. Furthermore, data is collected in different ways depending on the country and even within a single country, which leads to incomparable data and makes it difficult to conduct an objective analysis of global data [3, 5]. The availability of additional indicators derived from routinely collected data could improve the timeliness of epidemic outbreak detection.

Electronic health record (EHR) systems automatically collect a wide range of clinical data on patients throughout the healthcare process: medical history, vital signs, test orders and results, medical prescriptions, demographic information, and other data. EHR systems thus accelerate data transmission to epidemiological surveillance systems, reduce the workload on staff by eliminating the need for manual report submission, and improve the completeness of data available for monitoring. The system’s algorithms also accelerate disease detection prior to the definitive diagnosis required by traditional systems and can, among other things, help distinguish acute forms of disease from chronic ones based on testing history (e.g., hepatitis C) or identify conditions that lack reliable laboratory confirmation (e.g., culture-negative tuberculosis) [8–13]. However, the signals used in such systems for individual-level diagnosis may also reflect the state of the population. For example, inflammatory markers may indicate an increase in the prevalence of infectious diseases.

Coronavirus disease 2019 (COVID-19) serves as a useful model for studying this approach, as it is a well-studied disease caused by a novel pathogen for which the transmission routes, clinical manifestations, and mechanism of the immune response are known [14–17]. There are also highly sophisticated epidemiological models for COVID-19, such as Covasim [18]. We selected C-reactive protein (CRP) as our model object. It is a plasma protein routinely measured that is synthesized in the liver and serves as a sensitive biomarker of inflammation and tissue damage in acute infections, systemic inflammatory diseases, cardiovascular diseases, and cancers. CRP is not specific to any particular disease and responds more strongly to acute bacterial infections than to viral ones. Nevertheless, it is used as a biomarker of COVID-19 severity and exhibits rapid kinetics [19–24]. For this reason, CRP can serve as one of the indicators of an infectious disease outbreak in public health surveillance, particularly for new pathogens that do not yet have specific diagnostic tests, as was the case with COVID-19 [25, 26].

Although CRP levels rise sharply in COVID-19, the conditions under which an outbreak can be detected at the population level remain unclear. This question can be reliably investigated under controlled conditions using synthetic data from a model in which only the target pathogen with known dynamics is present, and other causes of elevated CRP are ruled out. An agent-based model (ABM) makes it possible to use the course of the disease for each individual to construct individual CRP dynamics and then aggregate this data to obtain a signal similar to the real one and assess detection capabilities in the absence of external factors.

However, in reality, at the population level, we are never dealing with a single concrete new pathogen, and CRP itself is not specific to infectious diseases, so detection using synthetic data provides only a proof of concept. Studying real-world routine laboratory tests during periods of seasonal infectious disease activity allows us to verify the reproducibility of the CRP patterns identified in the model and to assess the quality of detection under real-world conditions with known timelines of infection growth.

In this study, we investigated the possibility of detecting an increase in infectious disease activity based on routinely collected laboratory blood test data (Fig 1). First, we developed the BloodCovasim model, which simulates routine blood tests and individual CRP dynamics based on the disease progression of the agent in the Covasim epidemiological model [18]. Based on the simulation data, we determined the conditions under which an outbreak of the SARS-CoV-2 wild variant could be detected. Second, we confirmed the reproducibility of the model’s results using real-world data from routine blood tests during a period of seasonal increase in infectious activity in the summer and fall of 2025. We benchmarked different approaches to epidemic detection using both simulated and real-world data. Finally, we compared the reliability and sensitivity of using CRP with those of composite biomarkers.

**Fig 1.**
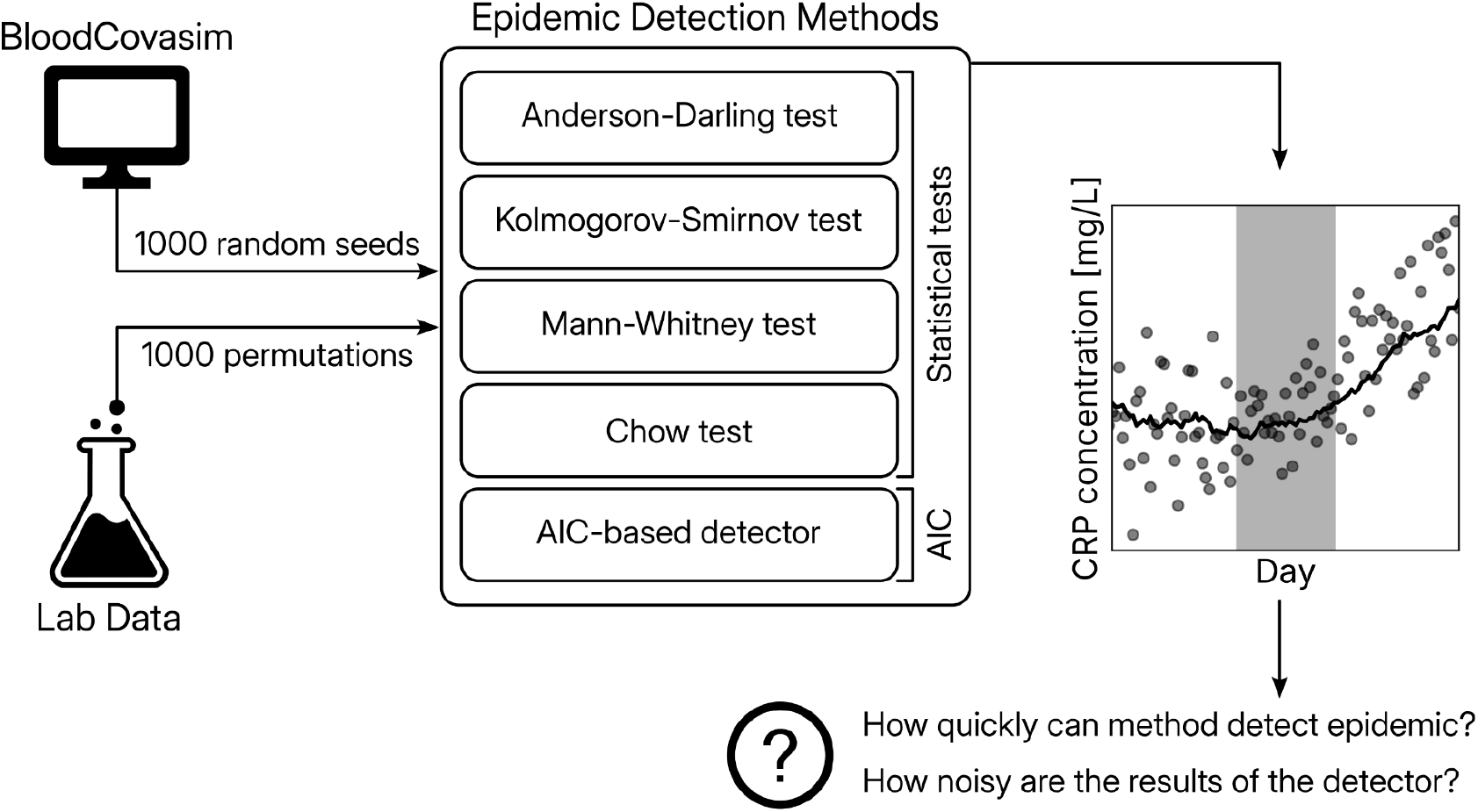
Description of the study flow.

## Methods

### Covasim

Covasim [18] is an agent-based model of the COVID-19 epidemic that describes the process at the level of individual agents (people). It simulates the personal course of the disease for each agent based on their age and their interactions with other people in the population (in households, at workplaces, in schools, and through community contacts) and allows researchers to study the course of the epidemic depending on the interventions implemented.

### BloodCovasim

To study the dynamics of CRP in a population during an epidemic, we implemented BloodCovasim. BloodCovasim handles the process of generating individual CRP dynamics and conducting blood tests in the population based on the results of the Covasim simulation.

For each agent, the baseline CRP value, in the absence of an epidemic agent in the body, is sampled from a lognormal distribution with a median of 4 mg/L and a 95% interval of [0.83 mg/L, 19.19 mg/L] (estimated from [27–31]). After determining the baseline CRP value, a parameter dynamics is generated. If the disease is symptomatic, then from the onset of symptoms until the day the most severe condition occurs (for a mild course of the disease, the increase occurs over 1 day), the CRP value increases linearly by an amount that depends on that condition and is sampled from a lognormal distribution with parameters from Table 1. CRP then decreases linearly until recovery to the agent’s baseline value. If the agent ultimately dies, their individual CRP trajectory is terminated, and they can no longer participate in the testing.

**Table 1.** Distribution parameters of peak CRP levels in patients with the disease in BloodCovasim, as a function of peak severity.

| Peak severity | Median peak CRP, mg/L | 95% peak CRP interval, mg/L | Sources |
| --- | --- | --- | --- |
| Mild | 12 | [4.50, 31.97] | [32] |
| Severe | 50 | [27.77, 90.02] | [31, 32] |
| Critical | 150 | [111.79, 201.27] | [28] |
| Dead | 250 | [205.50, 304.13] | [33] |

For ease of interpretation, the distributions were reported in terms of their median concentrations and central 95% intervals on the original CRP scale, rather than the mean and standard deviation of the corresponding log-transformed values. For a lognormal distribution of the form log *X ∼N* (*µ, σ*^2^), we specified a median value equal to exp (*µ*) and a central 95% interval equal to [exp (*µ−* 1.96*σ*), exp (*µ* + 1.96*σ*)].

If the disease is asymptomatic or the individual is not affected by the epidemic, their CRP remains at baseline. Examples of the resulting trends are shown in Fig 2A. The raw individual time series are smoothed using a one-dimensional Gaussian filter with a standard deviation for the Gaussian kernel equal to 2.

**Fig 2.**
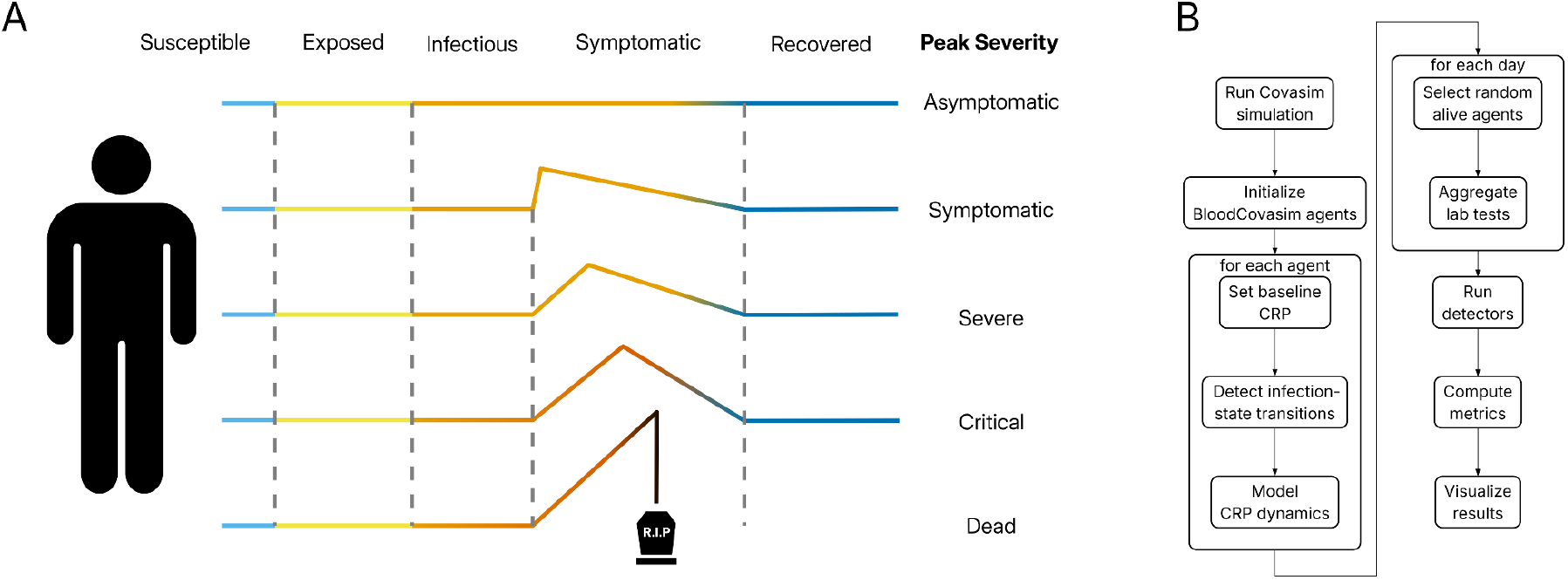
Generation and processing of synthetic CRP data in BloodCovasim. (A) A sample of individual CRP dynamics before smoothing, as a function of peak severity for a specific agent in BloodCovasim. (B) Flowchart representing processes for generating and processing synthetic data from BloodCovasim.

After initializing each agent, BloodCovasim collects data on individual CRP dynamics and then simulates random testing. During testing, for each simulation step, a specified number of agents that are alive at that moment are selected, and their CRP values are then aggregated (Fig 2B).

### Model validation

Kerr et al. [18] used data on the duration of various stages of the disease, the probabilities of disease progression, and relative susceptibility from COVID-19 studies, as well as real-world demographic parameters and artificial population structure parameters for various specific cities, when developing Covasim. In addition, the authors of the original paper validated Covasim using data on the number of infections, hospitalizations, ICU admissions, and deaths for the U.S. states of Washington and Oregon.

In developing BloodCovasim, we relied on studies of CRP and articles focusing specifically on CRP as a marker of disease progression in COVID-19 [27–33], which served as the basis for estimating the BloodCovasim parameters presented in Table 1. We also validated the internal algorithms underlying BloodCovasim and the study protocol.

### Design of experiments

Covasim simulations were run on a default artificial population of 100,000 agents for a duration of 300 steps (days), with 1,000 runs for each scenario. We varied the number of tests per day (10, 50, 250, and 1,250) to simulate different test coverage. In each run, the infectious agent was the SARS-CoV-2 wild variant, which was introduced into the population on day 100 in the form of 30 randomly selected infected agents. We disabled the possibility of reinfection because Covasim retains only the course of the most recent illness, which would have made the correct use of BloodCovasim impossible.

### Ethics Statement

The study involved a secondary analysis of retrospective routine laboratory data that had been de-identified before being provided to the authors. The authors had no access to information that could identify individual patients. The manuscript was reviewed by the institutional Ethics Committee and approved for publication on October 22, 2026. Given the retrospective nature of the study and the use of de-identified data, individual informed consent was not obtained.

### Real-world data preprocessing

We used data from 42,123 routine CRP and 202,218 complete blood count (CBC) tests from a commercial laboratory network in Russia for the period from July 1, 2025, to October 1, 2025. The data were filtered by age (0 to 85 years) and by percentiles (0.01% to 99.99%). Data from patients who were tested more than twice a year were also excluded.

### Calculation of inflammatory biomarkers

In our study, the traditional non-compound inflammatory biomarker CRP was compared with composite biomarkers [34–38] based on CBC tests, calculated as follows:

1. Systemic Inflammation Response Index (SIRI) = neutrophil count *×*monocyte count / lymphocyte count;
2. Systemic Immune-Inflammation Index (SII) = neutrophil count *×* platelet count / lymphocyte count;
3. Aggregate Index of Systemic Inflammation (AISI) = neutrophil count *×*monocyte count *×*platelet count / lymphocyte count;
4. Neutrophil-to-lymphocyte ratio (NLR) = neutrophil count / lymphocyte count;
5. Platelet-to-lymphocyte ratio (PLR) = platelets count / lymphocyte count;
6. Neutrophil-to-monocyte ratio (NMR) = neutrophil count / monocyte count. 0.05 *×* 10^9^*/*L was added to all denominators to prevent them from becoming 0.

### Outbreak detectors

This study involved the use of 5 different epidemic detectors. Throughout this manuscript, the term “detector” refers to a statistical method used to identify the epidemic outbreak from routinely collected blood test data.

The first group of detectors analyzed samples of values and compared them in consecutive 14-day windows using various basic statistical tests: the non-parametric Anderson-Darling test, the Kolmogorov-Smirnov test, and the Mann-Whitney test. A p-value less than 0.05, provided that the mean value increased between the samples under consideration, was considered a detector trigger corresponding to the first day following the observed values.

The second group of detectors used a moving average of the values over the previous 14 days. The first of these detectors calculated the Akaike information criterion (AIC) for constant, linear, and exponential models fitted to the 28-day moving average data and considered a trigger to have occurred if the minimum AIC was observed for the linear or exponential model, provided that the moving average over the last 14 days had increased compared to the previous 14 days (Fig 3). The second detector in this group compared the moving averages in consecutive 14-day windows using the Chow test to determine the presence of a structural break. The Chow test [39] tests the hypothesis that the coefficients of the linear regressions describing the data in consecutive windows are equal. A p-value below the 0.05 threshold, when the moving average over the last 14 days increased compared to the previous 14 days, was considered a trigger for the detector based on the Chow test. Triggers from these detectors were also assigned to the first day following the period under consideration.

**Fig 3.**
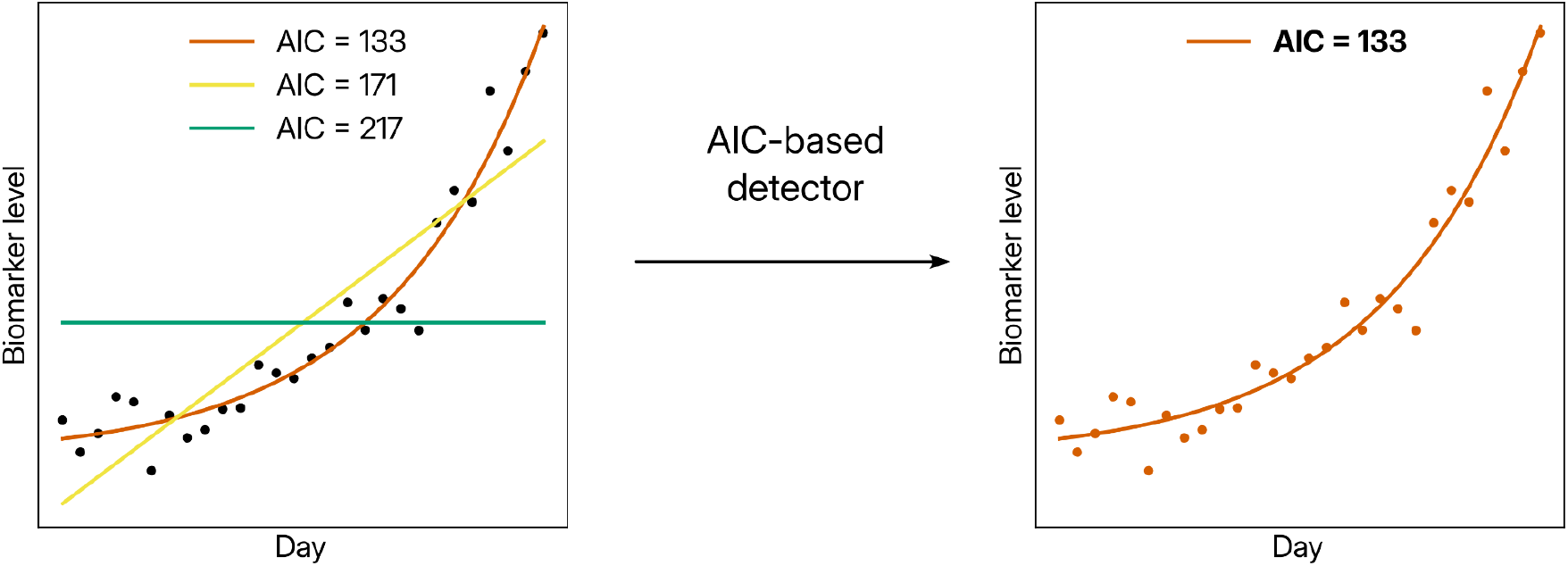
Scheme of the detection algorithm based on AIC. Constant, linear, and exponential models were fitted to 28 consecutive days of moving average of the studied values. The minimum AIC determined the best of the compared models for describing the data. A linear or exponential trend, in conjunction with an increase in the moving average, was considered a trigger for the AIC-based detector.

### Evaluation metrics

For synthetic data, the epidemic period (true positive) was defined as the interval between the day when 100 new daily infections were reached and the peak of new infections, shifted by 7 days to account for the incubation period. The interval prior to this was considered a non-epidemic period (true negative), and its start was shifted by 42 days (14 days for the moving average and 28 for the detectors) relative to the first day of the simulation so that all detectors would receive a complete dataset.

For the real-world data, the first two weeks of September 2025 were selected as the “true positive” period, as this is when the school year begins in Russia and the associated back-to-school effect occurs, contributing to a seasonal spike in the incidence of rhinoviruses, adenoviruses, seasonal coronaviruses, etc. [40–44]. To generate a set of samples based on the available data, we used bootstrapping, in which a specified number of values was selected pseudo-randomly with replacement for each day. True negatives were generated in a similar manner, but by sampling data from the true positive set, without any day-specific grouping, from the bootstrap sample as many times as necessary to ensure there were a sufficient number of days with data for the detectors by September 1, 2025. This allowed us to eliminate temporal correlations from the data, enabling us to assume no growth during the period under consideration. The data were collected starting July 1, 2025, to ensure that all detectors were functioning properly.

The target metrics were the peak detection rate in the true positive window, the ratio of the peak detection rate in the true positive window to that in the true negative window (signal-to-noise ratio, SNR), and the date relative to the start of the true positive window, when the detector first reached a 50 percent detection rate.

## Results

### Detection of an epidemic outbreak based on synthetic CRP test data

First, we investigated which methods are optimal to detect an epidemic in a simulation. We varied the number of daily tests (10, 50, 250, and 1,250) to reproduce different testing coverage. Each scenario was run in the BloodCovasim 1,000 times.

We found that when testing coverage is high (e.g., 1,250 daily tests), all methods perform approximately equally well (Fig 4D) and detect an increase in the number of infections in all model runs. When the number of tests is low, basic statistical tests detect an increase in incidence in 10-20% of runs, while moving average tests (AIC, Chow-based) detect it in 70-90% of runs (Fig 4C). The Kolmogorov-Smirnov detector is most affected by the low number of tests (e.g., 10 daily tests), with a peak detection rate of 13.1%, followed by the Anderson-Darling detector at 16.1% and the Mann-Whitney detector at 16.4%. Detectors based on moving averages prove to be more effective in this case, with a detection rate of 72.7% for the Chow test and 87.0% for the AIC (S1 Table).

**Fig 4.**
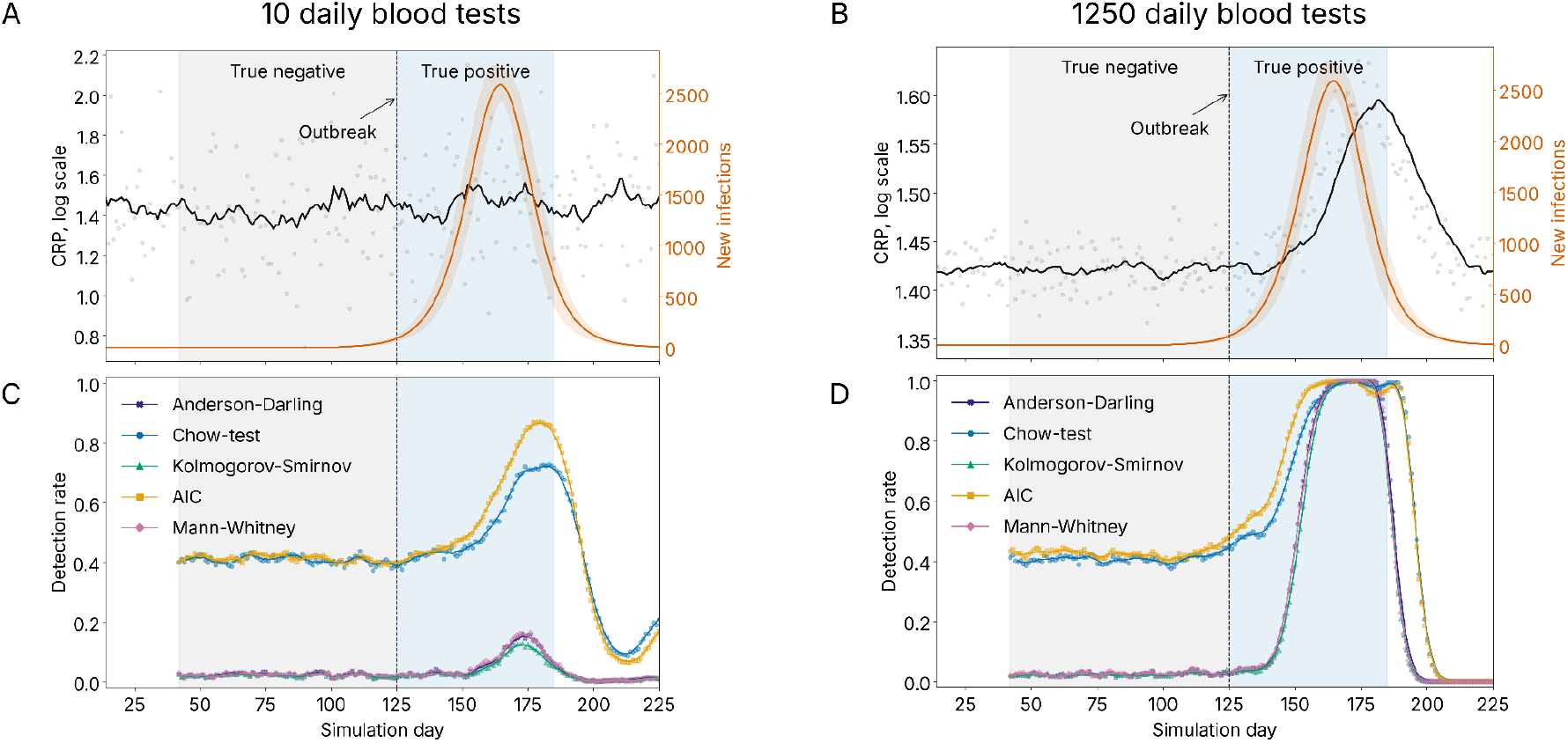
Outbreak detection results on synthetic CRP blood test data from BloodCovasim across 1000 different random seeds. CRP test data (black: data points represent the daily average log CRP for a single random seed; curve represents the moving average of log CRP over a 14-day window for the same seed) and new cases (orange: curve represents the average across random seeds; shading represents the standard deviation) for (A) 10 and (B) 1,250 daily blood tests. Detection rate for the detectors under study (data points: average detection probability on a selected day per random seed (detection rate); curve: 7-day centered moving average of the detection rate) for (C) 10 and (D) 1,250 daily blood tests. The gray area represents the days we consider to be the baseline, and the blue area represents what we consider to be the period of CRP increase.

Detectors using moving averages respond to the outbreak earlier (with AIC being slightly faster). The AIC-based detector reaches a 50% detection rate 3 days after the outbreak, the Chow test after 13 days, and the Anderson-Darling, Mann-Whitney, and Kolmogorov-Smirnov tests after 26-27 days, with 1,250 daily tests. At 10 daily tests, detectors based on basic statistical tests do not reach a 50% detection rate, while the AIC detector reaches it in 29 days and Chow in 35 (S1 Table).

When the mean CRP declines, moving-average-based detectors lag behind detectors based on basic tests by approximately two weeks (Fig 4D), which is a consequence of their use of a two-week moving average.

The detector based on the Chow test and AIC has a significantly higher level of background noise (false positives prior to an outbreak) than detectors based on basic statistical tests. At the same time, this background noise is practically independent of the number of daily tests (Fig 4C,D).

### Detection of seasonal increases in CRP due to a rise in the number of infectious diseases associated with the back-to-school effect

To compare different methods of epidemic detection in real-world settings, we analyzed CRP measurement results from a commercial laboratory network between July and September 2025. The expected increase in CRP levels is due to a rise in the prevalence of infectious diseases associated with increased contact density in early September (the back-to-school effect [40–44]). Data from July-August served as the baseline, while data from September served as the test data. A bootstrap analysis with measurement replacement was performed, and detector activations were evaluated in each permutation.

Based on real-world CRP data collected prior to the outbreak of infections associated with the start of the school year, the noise level in detectors using moving averages (AIC, Chow) is close to that observed in basic statistical tests (Fig 5). At the same time, AIC shows an increase in the detection rate approximately 7 days earlier than the others and reaches a detection rate of 85.3% (Fig 5B). Anderson-Darling reaches a peak detection rate of 81.4%, Mann-Whitney 71.3%, Kolmogorov-Smirnov 64.0%, and Chow 48.7%, at a testing rate of 250 tests per day (S1 Table). A rise in the Chow detection rate is also observed around September 26, 2025, caused by the moving average of the CRP reaching a plateau at the end of the true positive period.

**Fig 5.**
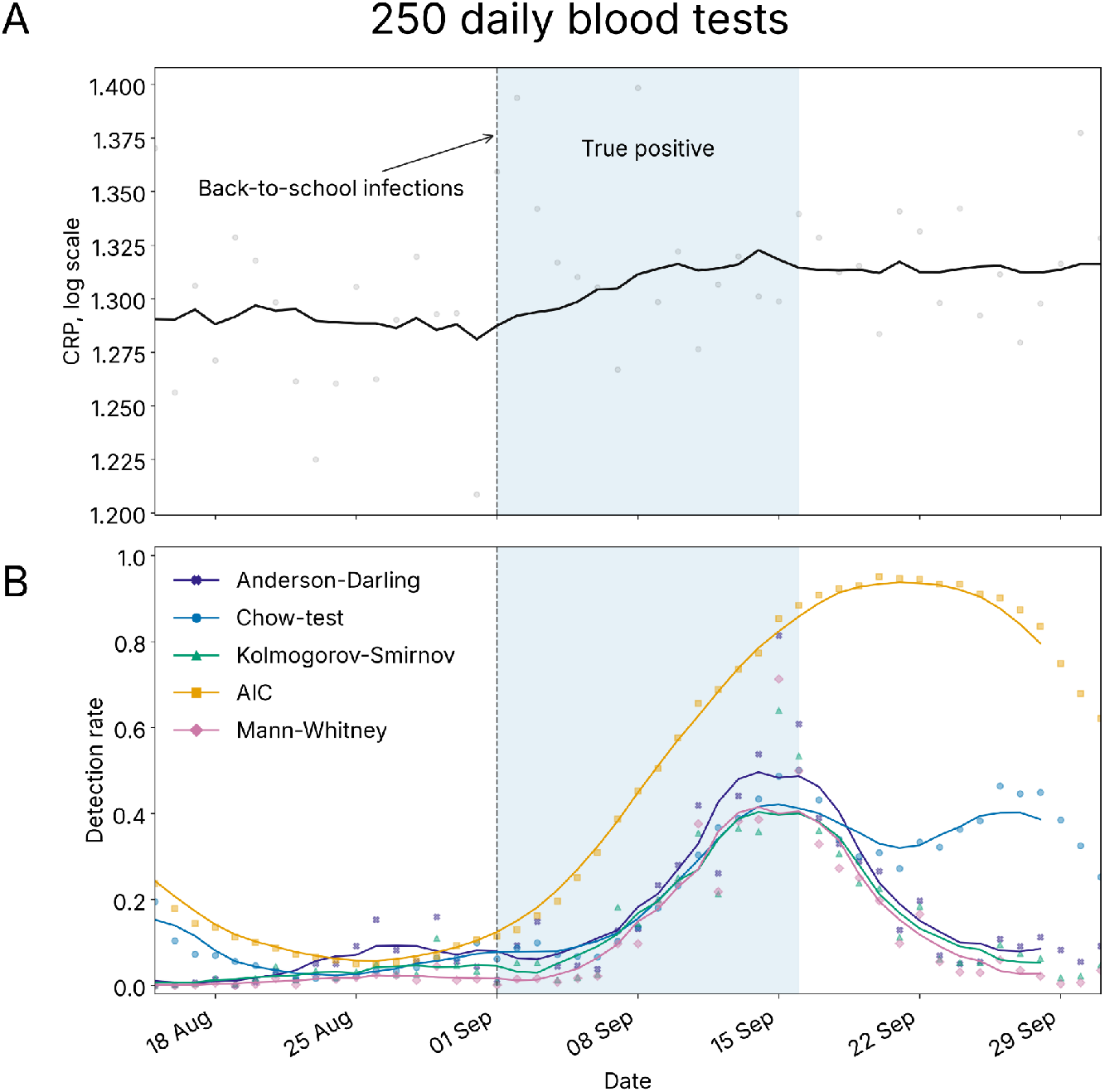
Results of back-to-school infection detection based on real-world CRP blood test data from Russia. CRP test data (black: data points represent the daily average log CRP; the curve represents the 14-day moving average of log CRP) for (A) 250 daily blood tests. Detection rate for the detectors under study based on 1,000 bootstrap samples (data points: average detection probability on a given day for a random seed (detection rate); curve: the 7-day centered moving average of the bootstrap detection probability) for (B) 250 daily blood tests. The blue area represents the first two weeks of September 2025, when we expect an increase in infectious diseases due to the back-to-school effect.

**Fig 6.**
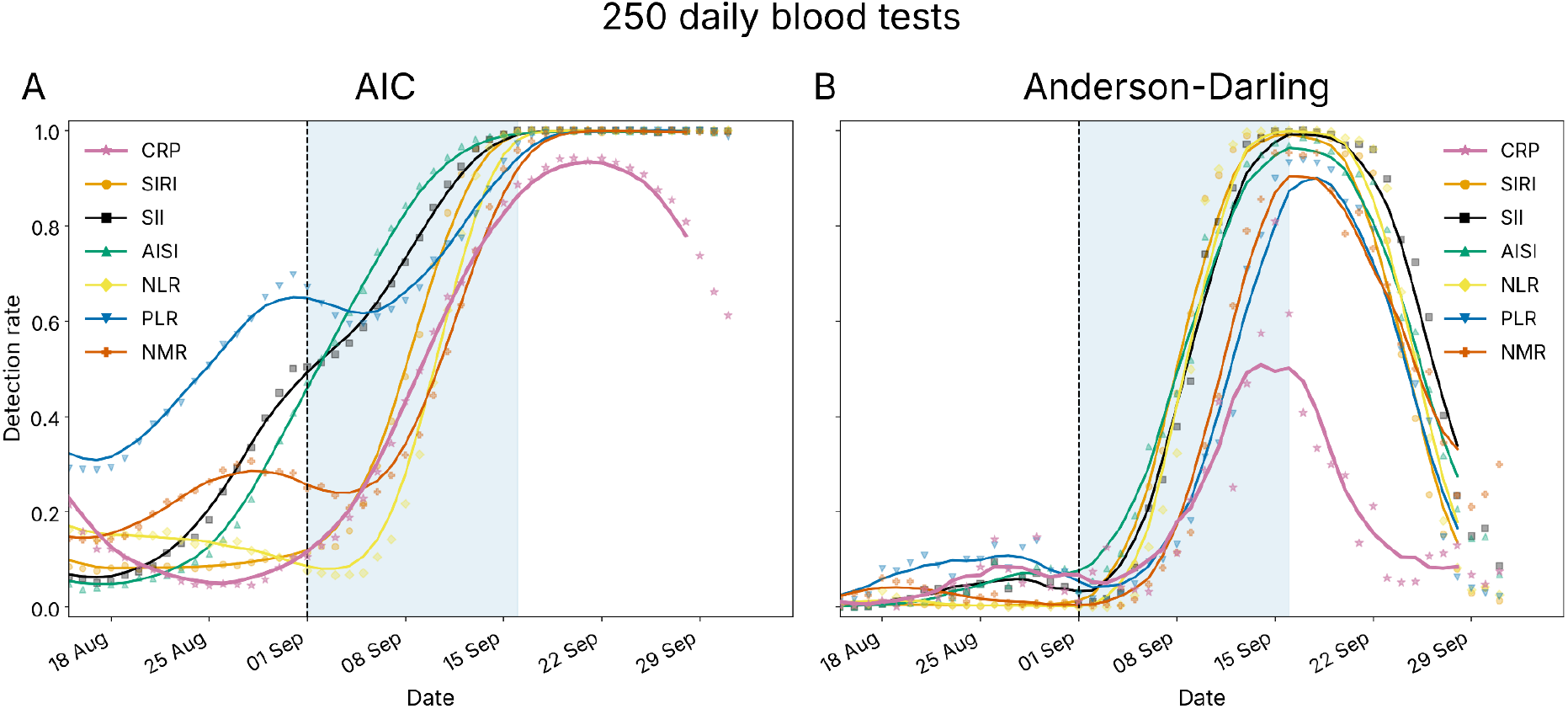
Results of back-to-school infection detection based on real-world CBC test data from Russia. Detection rates (A) for AIC and (B) Anderson-Darling detectors for 1,000 bootstrap samples of 250 daily blood tests (dots represent the average detection rate on a given day based on a random seed; the curve represents the 7-day centered moving average of the bootstrap detection probability) depending on the inflammation index used (SIRI, Systemic Inflammation Response Index = neutrophil count *×*monocyte count / lymphocyte count; SII, Systemic Immune-Inflammation Index = neutrophil count*×* platelet count / lymphocyte count; AISI, Aggregate Index of Systemic Inflammation = neutrophil count *×*monocyte count *×*platelet count / lymphocyte count; NLR, neutrophil-to-lymphocyte ratio = neutrophil count / lymphocyte count; PLR, platelet-to-lymphocyte = platelets count / lymphocyte count; NMR, neutrophil-to-monocyte = neutrophil count / monocyte count).

To evaluate how the tests in our study perform as a function of the number of measurements, we also conducted experiments with 50 or 10 tests per day (subsampling).

Reducing the number of tests lowers the peak detection rate, most significantly for the basic statistical tests, and also increases the noise level in the detectors based on running averages (more so for AIC) (S2 Fig). Although AIC becomes noisier through September, it yields a peak detection rate 6.5 times higher, with a SNR that is only 1.8 times lower than detectors using baseline tests for 10 daily tests (51.6% vs 7.9% for the Anderson-Darling peak detection rate and 1.23 vs 2.26 for the Kolmogorov-Smirnov SNR, S1 Table).

### A comparison of inflammatory indicators as markers of seasonal disease outbreaks

We also compared the effectiveness of detecting seasonal increases in incidence based on routine blood parameters other than CRP. We used composite indices associated with inflammation, including the most commonly measured parameters as a part of a CBC test: neutrophil, platelet, monocyte, and lymphocyte counts.

For each number of daily tests considered (10, 50, 250), the AIC and Anderson-Darling detectors require more CRP data than the other biomarkers under consideration. The peak detection rate for CRP is 84.8% and 80.9%, respectively, although for the other biomarkers it is >91.9% and >89.9% with 250 daily tests (S1 Table).

At the same time, the AIC detects growth 7-9 days earlier based on SII, AISI, and PLR data than based on SIRI, NLR, NMR, and CRP. Anderson-Darling reacts about 3 days earlier when using SIRI, AISI, NLR, and SII than when using PLR and NMR, and 5 days earlier than CRP. In this scenario, using AIC with SII and AISI data is most effective (S1 Table).

The difference between CRP and other biomarkers becomes more apparent as the daily number of tests decreases. With 10 daily tests, the best biomarkers yield a peak detection rate for the Anderson-Darling that is twice that of CRP (NLR) and 1.2 times that of CRP (SII). However, only the AIC detector reaches a 50% detection rate, and it does so earliest based on SII and AISI data (in about 6 days vs. 9+ for the others).

## Conclusion and discussion

In this study, we compared different approaches for the early identification of the spread of infectious diseases based on data on markers of inflammation. This is important because traditional methods of epidemic detection rely on data on disease-specific incidence and mortality, which are prone to underestimation and time lags and may not be applicable in the case of new emerging infections. Data from tests such as CRP blood tests and CBC tests, however, are routine measurements and are automatically entered into the laboratories informational or hospitals EHR systems regardless of the patient’s condition. They have the potential to be used for detecting new and emerging infections that may elude testing methods such as PCR and antibody tests. Of special interest is emergence of slowly progressing disease with subtle symptoms, but which nonetheless has an inflammatory component. That said, the methodology for use of data like routine blood test results for identifying epidemics remains an open question. As our study showed, the optimal method depends on the number of tests.

Using synthetic data from thousands runs of the BloodCovasim agent-based model and real-world data obtained from routine blood tests during the back-to-school period, we investigated detectors which compare distributions (the nonparametric Anderson-Darling test, the Kolmogorov-Smirnov test, and the Mann-Whitney test), as well as detectors based on moving averages (AIC-based and Chow test-based detectors).

We observed that detectors based on moving averages required less data (they have a higher probability of triggering with fewer tests), but exhibited higher noise levels when there is no trend in the data, and this noise was independent of the number of tests.

On synthetic data, the detectors demonstrate a lower data requirement (compared to real-world data) and, with the same number of daily tests (50, 250), achieve a detection rate of more than 80% (with a noise level of about 40%). Although detectors based on moving averages demonstrate a similar data requirement, AIC performs slightly better. This may be explained by the greater number of sources of variability in real-world data, for example, inflammation can result from trauma or other non-infectious diseases, which also exhibit seasonal patterns.

Based on real-world data, the AIC detects an increase in infections approximately one week earlier than other methods when 250 tests are conducted daily, and it has a higher probability of detection. When the number of daily tests decreases, it also outperforms other methods, exhibiting a significantly higher peak detection rate with a slightly lower signal-to-noise ratio.

CRP is not the only inflammatory biomarker. Even though it is convenient for modeling COVID-19 due to the availability of relevant studies [28, 31–33] and the fact that it is a non-compound parameter, there is no guarantee that CRP is the most reliable indicator of an increase in the number of infectious diseases.

Moreover, a CBC is the most commonly ordered test. Therefore, we compared CRP with composite biomarkers such as the Systemic Inflammation Response Index (SIRI), the Systemic Immune-Inflammation Index (SII), Aggregate Index of Systemic Inflammation (AISI), the neutrophil-to-lymphocyte ratio (NLR), the platelet-to-lymphocyte ratio (PLR), and the neutrophil-to-monocyte ratio (NMR) [34–38] as signals on real-world data for the same period as CRP.

Our analysis showed that, for any number of daily tests considered (10, 50, and 250), composite biomarkers outperform CRP in terms of sensitivity. In terms of timeliness, however, while not all outperform CRP, the best ones do. With 250 daily tests, the SII and AISI methods yielded the best results for the AIC-based detector, outperforming other biomarkers, CRP, and the Anderson-Darling test-based detector. As the number of tests decreases, the same biomarkers remain among the leaders for AIC, however, their advantage over CRP becomes even more pronounced, while the Anderson-Darling test-based detector no longer achieves a 50% peak detection rate for any biomarker.

However, the study has some limitations. BloodCovasim uses a highly simplified method for simulating individual CRP dynamics, based on only two reference points (baseline and peak) for each individual, due to a lack of data. In addition, testing in the real world occurs unevenly, the worse a person’s condition, the more likely they are to be tested. Failure to account for this could have underestimated the average CRP values and underestimated the effectiveness of the detectors when considering a scenario involving an outbreak of a new pathogen.

We also disabled the possibility of reinfection in the model because Covasim only stores the course of the agent’s most recent illness. However, in reality, reinfection with the SARS-CoV-2 wild variant is extremely rare (the reinfection rate for the wild variant is 0.26% [45]). The mere presence of reinfections affects the magnitude of the epidemic peak and the dynamics of the decline in the number of new cases at the end of the epidemic wave [46], but in this study we are primarily interested in outbreak detection and the growth period of new cases. Therefore, the absence of reinfections in the model does not distort the conclusions drawn.

Observations on different inflammation biomarkers may be pathogen-specific and relevant for viruses prevalent in early September in Russia (rhinoviruses, adenoviruses, RSV, seasonal coronaviruses). In general, it is recommended to use different inflammation biomarkers in surveillance as we do not know how a new emerging disease will develop.

Thus, we have demonstrated that population-based blood tests for CRP and CBC can serve as markers of an epidemic outbreak, whether it involves seasonal illnesses such as “back-to-school” infections or new pathogens such as the SARS-CoV-2 wild variant. Our study may help epidemiologists select the biomarkers and detectors to use in epidemic surveillance, depending on the volume of incoming data. A basic recommendation is to use detectors based on the AIC when using moving averages and on the

Anderson-Darling test, as it is the most sensitive test among the basic statistical methods.

## Data Availability

All code used to generate and analyze the synthetic data and to analyze the real-world data, together with the resulting aggregated metrics and figure-generation scripts, is publicly available at https://github.com/HHDM-SBM/bloodcovasim-paper. The synthetic datasets can be fully regenerated from the provided code and parameters. Individual-level human laboratory data cannot be openly shared due to privacy restrictions but may be accessed by qualified researchers upon reasonable request and approval by the data provider.

## Supporting information

**S1 Fig.**
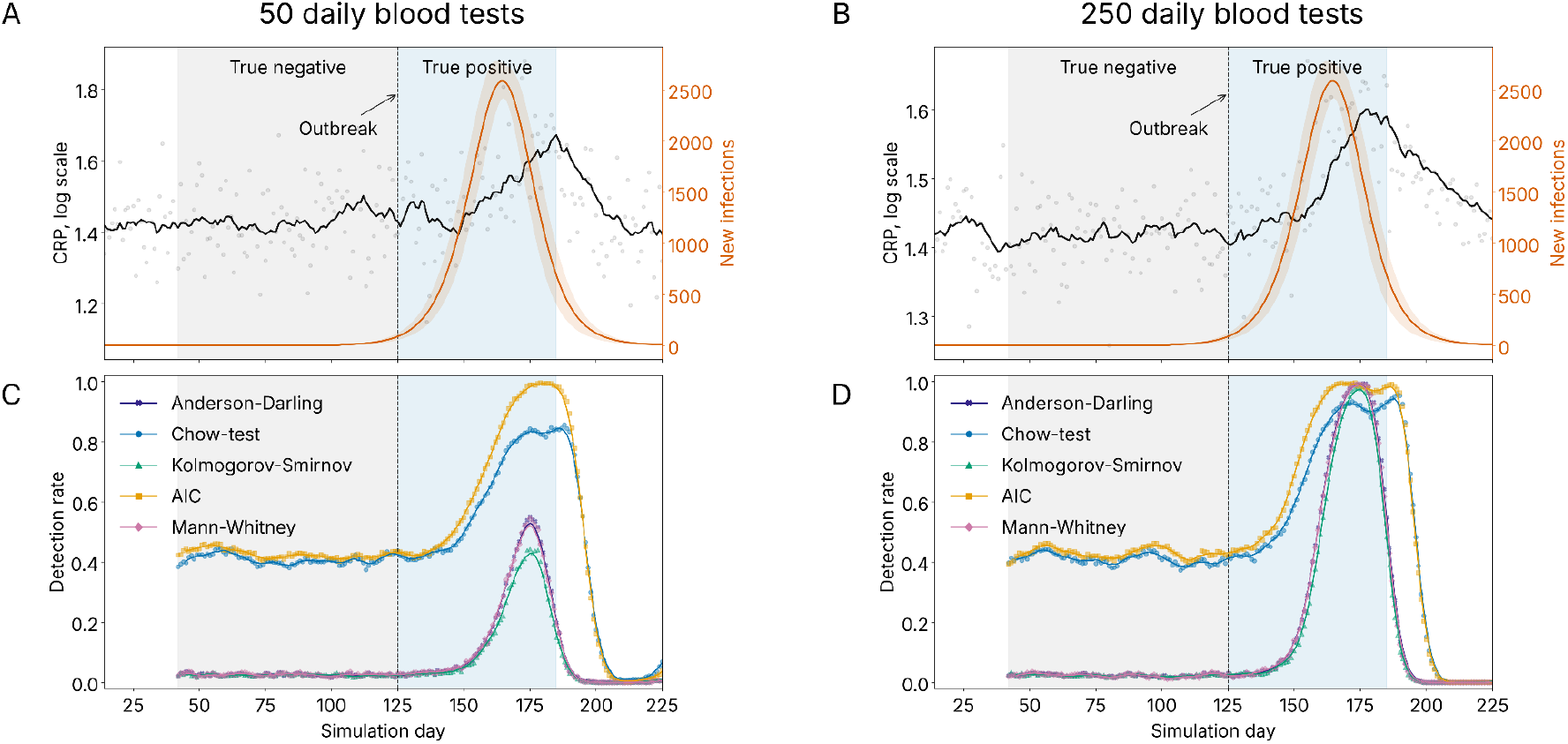
Outbreak detection results on synthetic CRP blood test data from BloodCovasim across 1000 different random seeds. CRP test data (black: data points represent the daily average log CRP for a single random seed; curve represents the moving average of log CRP over a 14-day window for the same seed) and new cases (orange: curve represents the average across random seeds; shading represents the standard deviation) for (A) 50 and (B) 250 daily blood tests. Detection rate for the detectors under study (data points: average detection probability on a selected day per random seed (detection rate); curve: 7-day centered moving average of the detection rate) for (C) 50 and (D) 250 daily blood tests. The gray area represents the days we consider to be the baseline, and the blue area represents what we consider to be the period of CRP increase.

**S2 Fig.**
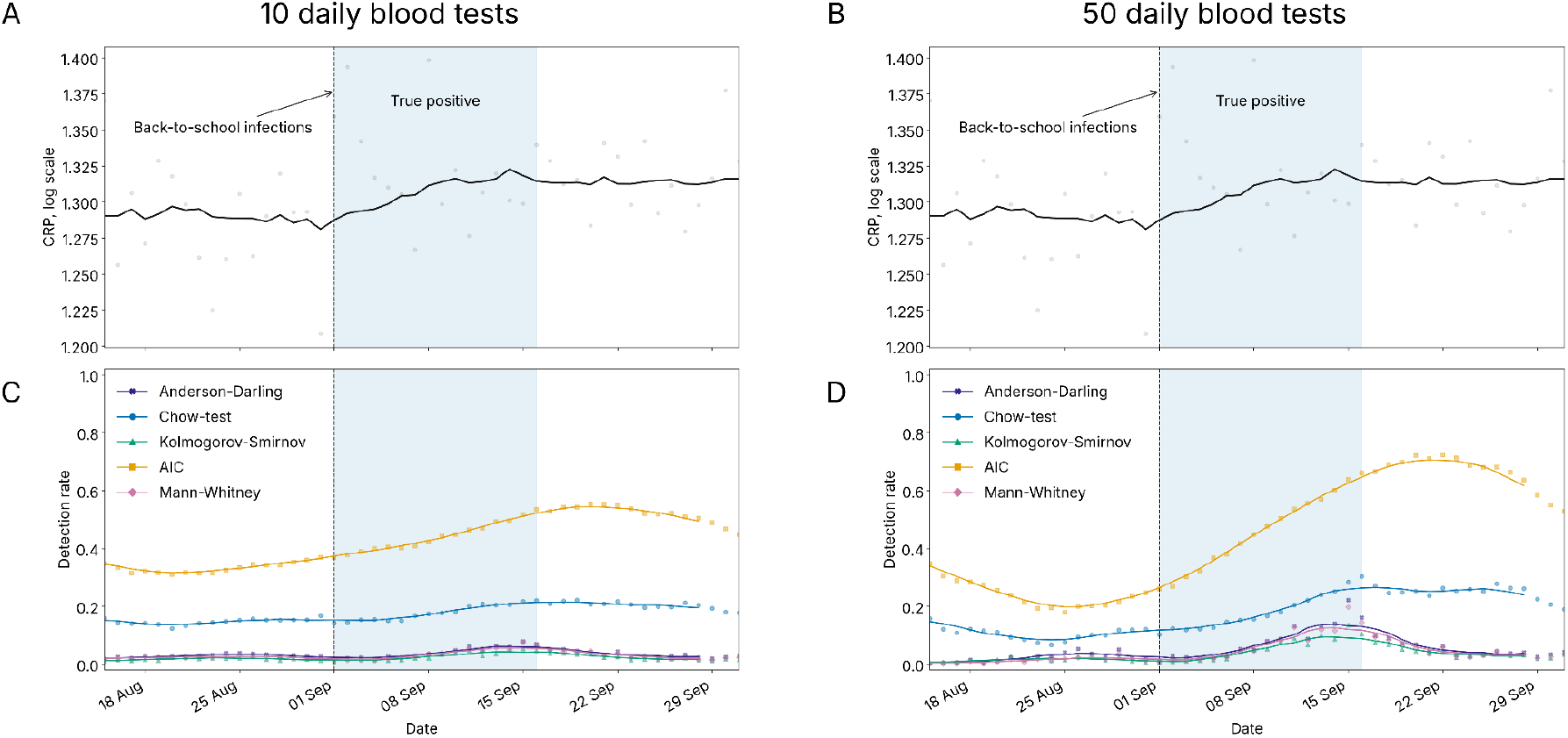
Results of back-to-school infection detection based on real-world CRP blood test data from Russia. CRP test data (black: data points represent the daily average log CRP; the curve represents the 14-day moving average of log CRP) for (A) 10 and (B) 50 daily blood tests. Detection rate for the detectors under study based on 1,000 bootstrap samples (data points: average detection probability on a given day for a random seed (detection rate); curve: the 7-day centered moving average of the bootstrap detection probability) for (C) 10 and (D) 50 daily blood tests. The blue area represents the first two weeks of September 2025, when we expect an increase in infectious diseases due to the back-to-school effect.

**S3 Fig.**
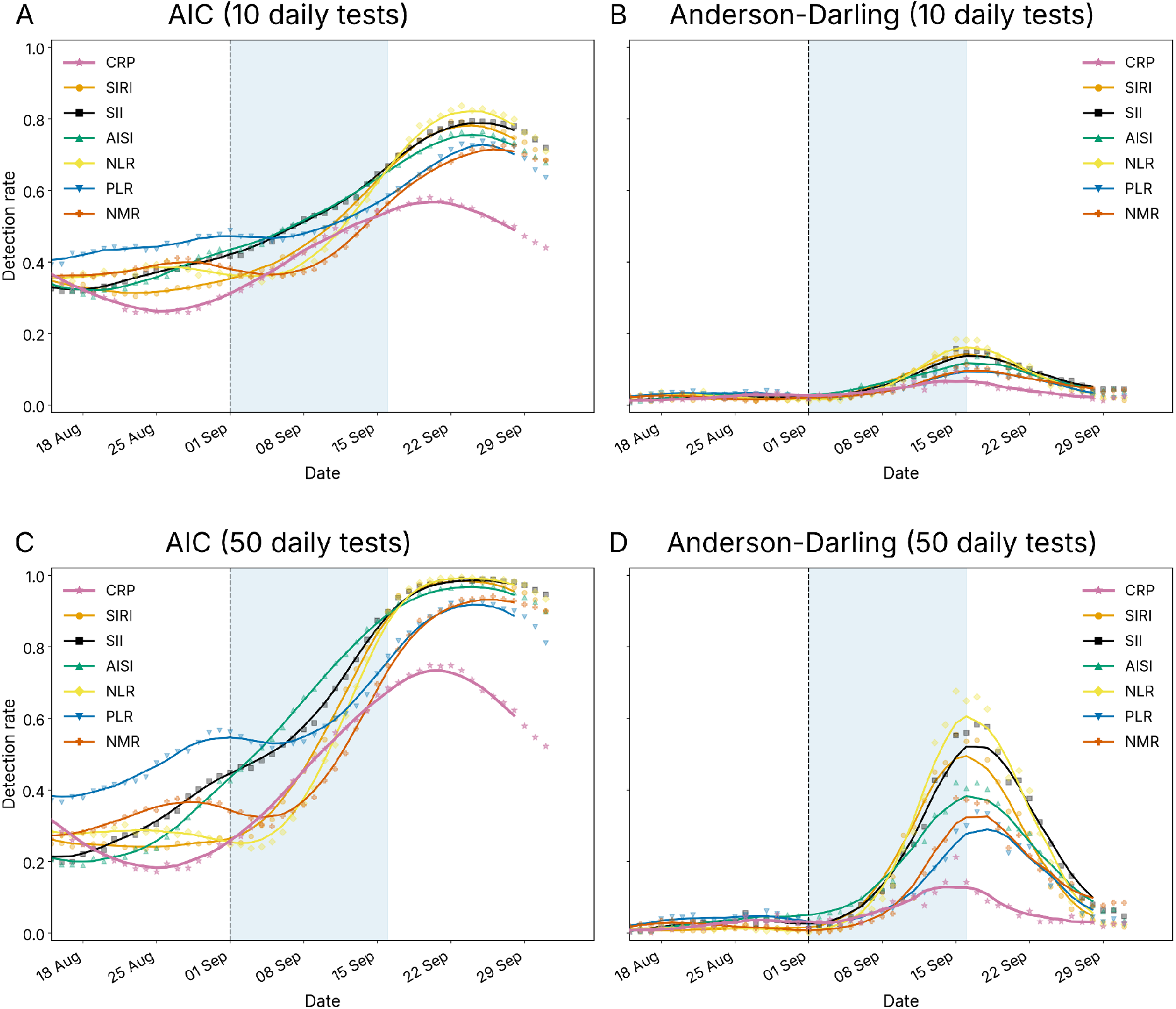
Results of back-to-school infection detection based on real-world CBC test data from Russia. Detection rates (A,C) for AIC and (B,D) Anderson-Darling detectors for 1,000 bootstrap samples of (A,B) 10 and (C,D) 50 daily blood tests (dots represent the average detection rate on a given day based on a random seed; the curve represents the 7-day centered moving average of the bootstrap detection probability) depending on the inflammation index used (SIRI, Systemic Inflammation Response Index = neutrophil count *×*monocyte count / lymphocyte count; SII, Systemic Immune-Inflammation Index = neutrophil count *×*platelet count / lymphocyte count; AISI, Aggregate Index of Systemic Inflammation = neutrophil count *×*monocyte count *×* platelet count / lymphocyte count; NLR, neutrophil-to-lymphocyte ratio = neutrophil count / lymphocyte count; PLR, platelet-to-lymphocyte = platelets count / lymphocyte count; NMR, neutrophil-to-monocyte = neutrophil count / monocyte count).

**S1 Table. Evaluation metrics for the detectors under study in all the experiments of our research.**

